# Prevalence of Non-Suicidal Self-Injury, Depression, and Anxiety among In-School Adolescents in Gboko LGA, Benue State, Nigeria

**DOI:** 10.1101/2025.09.25.25336377

**Authors:** Vershima Daniel Zar, Udo Winifred Mbarumun, Ochigbo Enenu, Ibrahim Umar, Wamanyi Yohanna, Emmanuel Etim Clement, Agbaje Samson

## Abstract

**Background:** Non-Suicidal Self-Injury (NSSI) is widely recognised in psychological literature as the deliberate infliction of harm upon one’s own body—such as cutting, hitting, or burning— without suicidal intent. This behaviour has garnered increasing attention due to its prevalence and the complex psychological factors underlying its manifestation. This study examined the baseline levels of NSSI, depression, and anxiety within the specific adolescent school population of Gboko LGA.

**Methods:** This study was conducted in Gboko Local Government Area (LGA) of Benue State, Nigeria, and employed a correlational research design to address its stated objectives. The target population comprised all secondary school students enrolled in Gboko LGA. Data were collected using the Predictors of Non-Suicidal Self-Injury Questionnaire (PNSSIQ) from a sample of 420 in-school adolescents. Data analysis was conducted using descriptive statistics. Research Question 1 was analysed using frequencies and percentages, while Research Questions 2 and 3 were examined using mean scores and standard deviations.

**Results:** Findings showed that in-school adolescents had a low proportion of engagement in NSSI behaviour. The finding was somewhat expected, considering the strong familial and community ties often observed in Nigerian cultural settings, which may provide protective support against such behaviours. Furthermore, the study showed that in-school adolescents had a mild level of anxiety in Gboko LGA, Benue State.

**Conclusion:** This study highlighted the urgent need for culturally sensitive mental health interventions, increased awareness, and destigmatization efforts to ensure accurate assessment and effective support for adolescents experiencing psychological distress. Conclusively, the study underscores the complex interplay between cultural context and adolescent mental health, and calls for targeted strategies that respect and leverage local values while addressing emerging mental health needs. It was recommended, amongst others, that public health interventions should focus on enhancing parental attachment and involvement in adolescents’ lives.

## Introduction

Non-suicidal self-injury (NSSI) or non-suicidal self-harm (NSSH) is a major public health problem among adolescents. Swannel, Martin, Page, Heskiry and John (2014) reported that NSSI has a lifetime prevalence of 17 per cent in adolescents and young adults (10-24 years) and 74 per cent in young adults with psychiatric disorders. Among Chinese students, the prevalence of NSSI ranged from 24.9 to 29.2 per cent (Wan et al., 2015; Tang et al., 2016). Furthermore, studies (Turecki & Brent, 2016; McManus et al., 2019) reported that NSSI is associated with suicidal attempt and death by suicide. World Health Organization (2022) reported that injuries due to both unintentional causes and violence claimed the lives of 4.4million people globally in 2019 and constituted eight per cent of all deaths.

The prevalence of NSSI seems to be high among adolescents and young adults in Sub-Saharan Africa (SSA). Khaled et al. (2024) reported a high 12-month prevalence estimate of 24·3 per cent (IQR = 16·9 per cent - 27·9per cent). Quarshie, Waterman and House (2020) reported that the median lifetime prevalence estimate of NSSI was 10·3 per cent (interquartile range [IQR] 4·6 per cent - 16·1 per cent); median 12-month prevalence estimate was 16·9 per cent (IQR: 11·5 per cent - 25·5 per cent); median 6-month prevalence estimate of NSSI was 18·2 per cent (IQR: 12·7 per cent - 21·8 per cent); and the median 1-month prevalence estimate of NSSI was 3·2 per cent (IQR: 2·5-14·8 per cent) among young adults in Sub-Saharan Africa. Quarshie et al. (2020) further reported that clinical samples of adolescents commonly reported overdose of drugs, whereas self-cutting was most reported in non-clinical samples.

Non-suicidal self-injury (NSSI) or self-harm (NSSH) is a behavioural problem among Nigerian adolescents. Azubuike and Onyemaka (2012) reported that the prevalence of self-injury was 73.6 per cent among adolescents in Agbor Metropolis, Delta State. Opakunle, Aloba, Suleiman and Akinsulone (2019) reported that the 12-month prevalence of suicidal ideation was 12 per cent among in-school adolescents in Osogbo, Osun State. Therefore, NSSI is a significant health problem among adolescents in Nigeria, including Benue State.

Non-suicidal self-injury (NSSI) has been conceptualised in the literature. Nock and Favazza (2009) defined NSSI as deliberate destruction of one’s own body (e.g., cutting, hitting or burning of the skin) in the absence of suicidal intent. It is often used interchangeably with self-harm, though it is important to bear in mind that self-harm may carry harmful and non-harmful consequences. The International Society for the Study of Self-Injury (ISSI, 2018) defined non-suicidal self-injury as the deliberate, self-inflicted damage of body tissue without suicidal intent and for purposes not socially or culturally sanctioned. This definition has several important parts: first, the harm that results from self-injury is an intentional or expected consequence of the behaviour; second, self-injury usually results in some sort of immediate physical injury, including cuts, bruises, scratches, or marks on the skin. Behaviours that do not directly result in injuries are usually excluded, even though they may be harmful or dangerous. Third, self-injury is separate from suicidal thoughts or behaviours, in which individuals want to end their lives. People usually report that they have no expectation or intention to cause death when they engage in self-injury (ISSI, 2018).

Though less frequent, NSSI is sometimes referred to by focusing on particular methods (e.g., self-cutting). While cutting is among the most widely recognized forms of self-injury, the behaviour can take many other forms, including burning, hitting, or scratching oneself. Furthermore, many people who self-injure report using more than one method during their lives (Whitlock et al., 2011). In this study, NSSI refers to the direct and intentional destruction of one’s own body tissues without the plan of killing oneself. Also, non-suicidal self-injury (NSSI) behaviour or non-suicidal self-harm (NSSH) behaviour will be used interchangeably in this study.

The various methods adolescents use to self-injure have been identified in literature. The methods include superficial to moderate self-cutting, burning, scratching, and bruising. Other examples of NSSI involve cutting, burning, stabbing, or excessive rubbing (Whitlock et al., 2011). The behaviours of NSSI are performed with the expectation that the injury produced will be minor to moderate and will not be life-threatening. However, behaviours such as nail biting and practices like piercing and tattooing are not included in the proposed definition for NSSI (Daniel, 2020).

The adverse effects of NSSI on adolescents’ health are numerous. Involvement in NSSI is strongly associated with many poor mental health outcomes, such as low self-esteem, depression, anxiety, and suicide attempts and psychiatric distress. Accordingly, it is unsurprising that the most common function of NSSI reported in non-clinical (i.e. community) samples of adolescents is regulation of difficult and intense thoughts and feelings (McManus et al., 2019). Adopting NSSI as a behaviour could be influenced by certain factors.

Some factors may trigger or initiate the occurrence of NSSI among adolescents, including in-school adolescents in Gboko LGA, Benue State. Such factors can be grouped into personal, parental, psychological, social and environmental factors (Nock & Favazza, 2009; Quershie et al., 2020). These factors are also known as predictors. Skagerstrom, Alehagem, Haggstrom-Nordin, Anestedt and Nilsen (2013) defined a predictor as an event or something that shows what will happen in the future. Predictor refers to something such as an event or fact that enables the explanation of what will happen in the future (Hornby, 2015). In this study, predictors refer to a factor that can influence or initiate the occurrence of NSSI among in-school adolescents. This study seeks to examine some factors of NSSI among in-school adolescents. Examples of predictors of NSSI behaviours include the environmental, social, cultural, psychological, familial, and institutional factors (O’Connor & Nock, 2014). However, in this study, the factors will be restricted to sociodemographic factors, psychological factors, and personal factors because they have more influence on the occurrence of NSSI among adolescents (Nock & Favazza, 2009; Quarshie et al., 2020; Aboagye et al., 2022). Psychological factors of interest include depression and anxiety. Personal factors include self-esteem and parental attachment.

### Statement of the Problem/Justification

Adolescents’ overall health and well-being are critical to their development, growth, survival and academic achievements. Adolescence also provides opportunities for adopting healthy lifestyles, embracing health-promoting behaviours, and achieving optimal mental health. Thus, maintaining optimal well-being may be a protective factor against non-suicidal self-injury (NSSI) among adolescents. However, adolescence, being a transition phase of life, is characterised by turbulence, experimentation and health risk behaviours. Young people, including in-school adolescents, are vulnerable to risk-taking due to emotional instability and desires to experiment with unhealthy behaviours and cope with unpleasant life events. Such coping measures include NSSI. Non-suicidal self-injury aims at inflicting harm on oneself with attendant health consequences. Such consequences include suicide, suicidal thoughts and psychiatric distress. Non-suicidal self-injuries can be triggered by certain factors. The factors can be personal, psychological, social and parental. The influences of these factors on in-school adolescents’ involvement in NSSI vary from one person to another and from one country to another. In addition, the literature evidence on the association between these factors and engagement in NSSI among adolescents is mixed and inconclusive. Therefore, the present study sought to determine the baseline levels of NSSI, depression, and anxiety of in-school adolescents’ involvement in NSSI. Ascertaining these factors can inform the design of appropriate public health interventions. The researcher, therefore, intends to determine predictors of NSSI among in-school adolescents in Gboko LGA, Benue State.

### Aim and Objectives of the Study

This study aimed to investigate the baseline levels of NSSI, depression, and anxiety within the specific adolescent school population of Gboko LGA. The following objectives guided the study:

1. To determine the proportion of in-school adolescents who engage in NSSI in Gboko LGA, Benue State.
2. To determine the level of depression among in-school adolescents in Gboko LGA, Benue State;
3. To find out the level of anxiety among in-school adolescents in Gboko LGA, Benue State;

### Research Questions

The following research questions were posed to guide the study:

1. What is the proportion of in-school adolescents who engage in NSSI in Gboko LGA, Benue State?
2. What is the level of depression among in-school adolescents in Gboko LGA, Benue State?
3. What is the level of anxiety among in-school adolescents in Gboko LGA, Benue State?

## Method and Materials

This study was conducted in Gboko Local Government Area (LGA) of Benue State, Nigeria, and employed a correlational research design to address its stated objectives. The target population comprised all secondary school students enrolled in Gboko LGA during the 2022/2023 academic session. Specifically, the population included 9,240 in-school adolescents, as reported by the Benue State Teaching Service Board, Makurdi (2023). A sample of 420 in-school adolescents was selected for the study. The sample size was determined using Yamane’s (1967) formula for sample size calculation, and a multi-stage sampling technique was employed to ensure representativeness. Data were collected using a structured questionnaire titled Predictors of Non-Suicidal Self-Injury Questionnaire (PNSSIQ). Embedded within the PNSSIQ was the Self-Harm Inventory (SHI), developed by Sansone, Wiederman, and Sansone (1998). The SHI is a 22-item self-report instrument designed to assess a wide spectrum of self-harming behaviours. The questionnaire also contained the 9-item version of the Patient Health Questionnaire (PHQ-9) and the GAD-7, which is a self-administered seven-item instrument used as a screening tool for anxiety. The PHQ-9 and GAD-7 were developed by Kroenke, Spitzer, and Williams (2001). Respondents rated each item on a five-point Likert scale ranging from 0 to 4, where 0 = “Disagree,” 1 = “Somewhat Disagree,” 2 = “Neither Agree nor Disagree,” 3 = “Somewhat Agree,” and 4 = “Agree.” The instrument underwent content validation by three subject-matter experts. Reliability was assessed using Cronbach’s alpha, yielding a coefficient of 0.84, indicating a high level of internal consistency.

Data collection was executed in phases. Before commencement, the researcher obtained formal permission from the administrative heads of the selected institutions. Three research assistants, comprising teachers and community health workers currently serving in Gboko LGA, were trained on the procedures for administering the instrument and retrieving completed questionnaires. The researcher and assistants visited the selected secondary schools on pre-arranged dates. A total of 420 questionnaires were distributed to students during their break periods. Participants were encouraged to complete the instrument immediately and return it to the researcher or assistants. For those unable to do so on the spot, completed questionnaires were submitted to class teachers for subsequent collection. Data analysis was conducted using descriptive statistics. Research Question 1 was analysed using frequencies and percentages, while Research Questions 2 and 3 were examined using mean scores and standard deviations.

## Results

Table 1 shows that most adolescents involved in the study were aged 15–19 years (49.2%), with slightly more females (54.7%) than males (45.4%). The majority were in junior secondary classes, particularly JSS2 (37.0%) and JSS1 (32.8%). Most came from medium-sized families (42.8%), resided in urban areas (59.0%), and had parents with no formal education (32.5%), followed by those with secondary (22.3%) and tertiary education (20.9%). Furthermore, most respondents attended combined schools (71.4%) compared to boys-only (13.3%) and girls-only schools (12.8%).

**Table 1:**
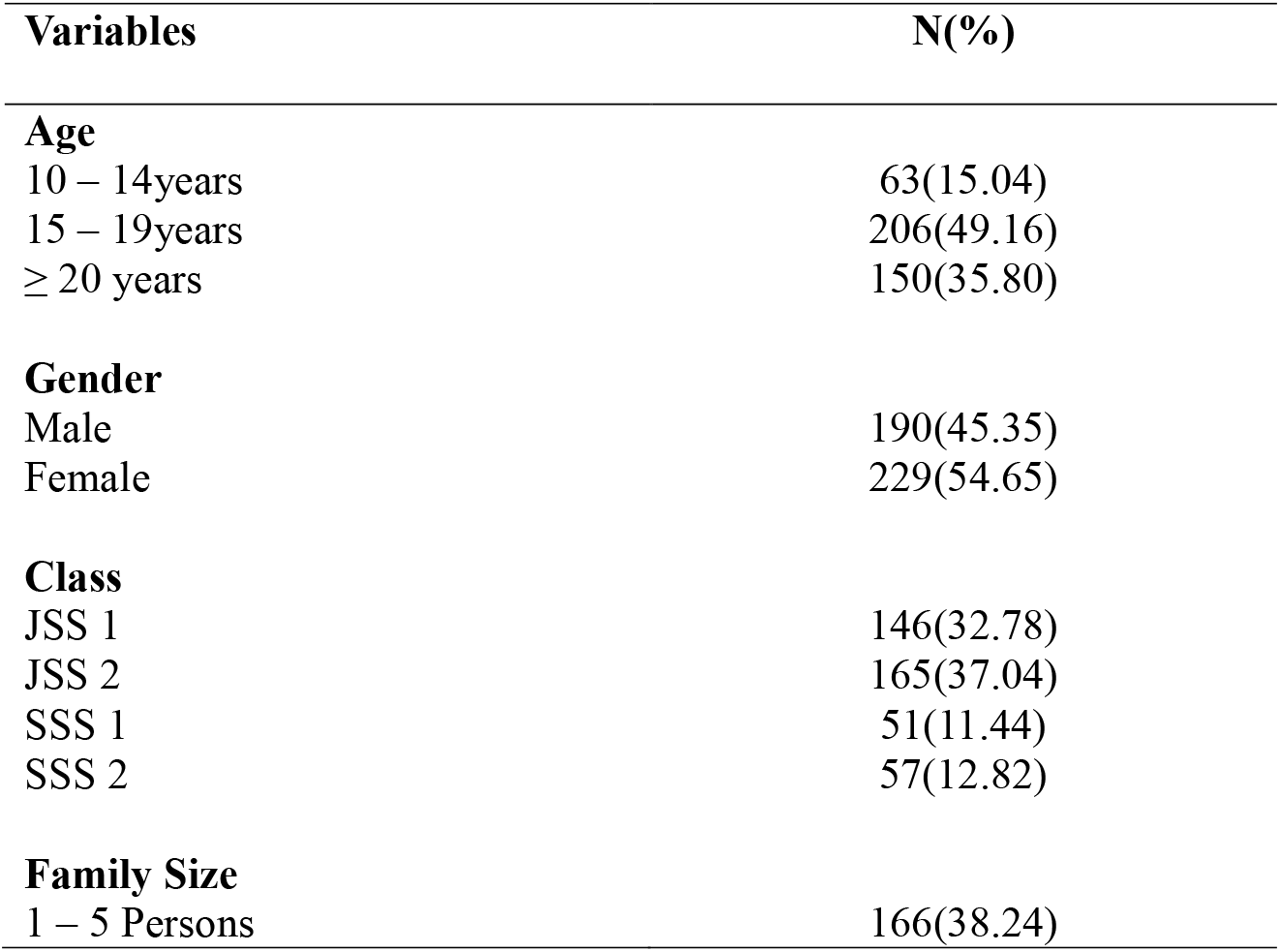
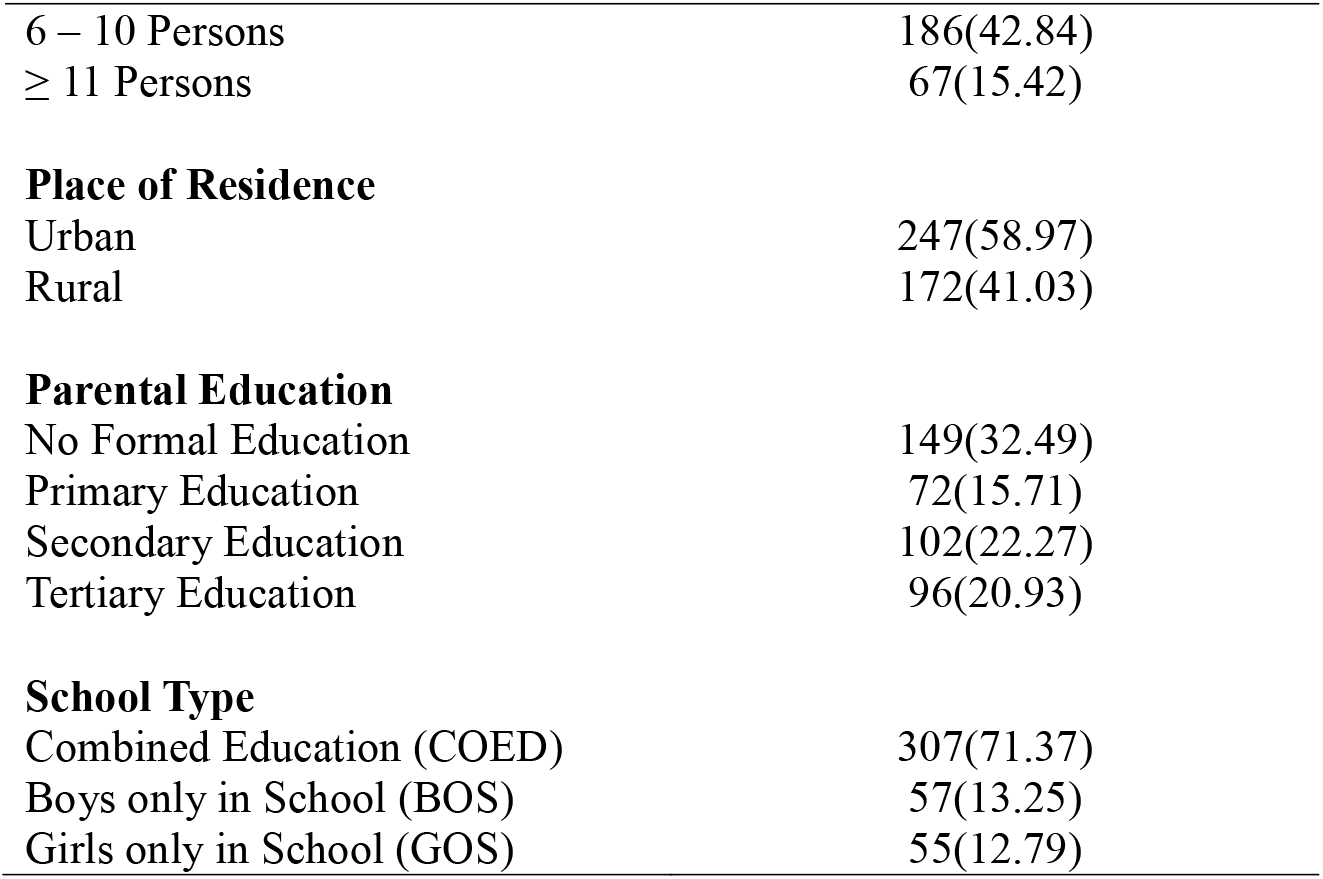
Sociodemographic Characteristics of Respondents (n-419)

| **Variables** | **N(%)** |
| --- | --- |
| **Age** |  |
| 10 – 14years | 63(15.04) |
| 15 – 19years | 206(49.16) |
| ≥ 20 years | 150(35.80) |
| **Gender** |  |
| Male | 190(45.35) |
| Female | 229(54.65) |
| **Class** |  |
| JSS 1 | 146(32.78) |
| JSS 2 | 165(37.04) |
| SSS 1 | 51(11.44) |
| SSS 2 | 57(12.82) |
| **Family Size** |  |
| 1 – 5 Persons | 166(38.24) |
| 6 – 10 Persons | 186(42.84) |
| ≥ 11 Persons | 67(15.42) |
| **Place of Residence** |  |
| Urban | 247(58.97) |
| Rural | 172(41.03) |
| **Parental Education** |  |
| No Formal Education | 149(32.49) |
| Primary Education | 72(15.71) |
| Secondary Education | 102(22.27) |
| Tertiary Education | 96(20.93) |
| **School Type** |  |
| Combined Education (COED) | 307(71.37) |
| Boys only in School (BOS) | 57(13.25) |
| Girls only in School (GOS) | 55(12.79) |

Table 2 presents the prevalence of non-suicidal self-injury (NSSI) behaviors among in-school adolescents in Gboko Local Government Area, Benue State (n = 419). The analysis showed that, overall, 11.4% of respondents reported engaging in at least one form of NSSI behavior, while 88.6% did not. Based on the classification scale (0–39% = low, 40–59% = moderate, 60–79% = high, 80% and above = very high), this indicates a low overall proportion of NSSI among the study population.

**Table 2:** Proportion of In-school Adolescents Who Engaged in NSSI Behaviours in Gboko LGA, Benue State (n = 419)

| S/N | SHI Items | Yes<br>f(%) | No<br>f(%) |
| --- | --- | --- | --- |
| 1. | Overdosed on drugs | 83(19.8) | 336(80.2) |
| 2. | Cut yourself on purpose? | 51(12.2) | 368(87.8) |
| 3. | Burned yourself on purpose | 33(7.9) | 386(92.1) |
| 4. | Hit yourself | 58(13.8) | 361(86.2) |
| 5. | Banged your head on purpose | 49(11.7) | 370(88.3) |
| 6. | Abused or drink too much alcohol? | 37(8.8) | 382(91.2) |
| 7. | Driven your father's or mother's car or bike recklessly on purpose | 58(13.8) | 361(86.2) |
| 8. | Scratched yourself on purpose | 47(11.2) | 372(88.8) |
| 9. | Prevented your wound from healing by not taking drugs or dressing it | 65(15.5) | 354(84.5) |
| 10. | Made medical situations worse on purpose (e.g., skipped prescribed Medication/drugs) | 63(15.0) | 356(85.0) |
| 11. | Having sex with many girlfriends or boyfriends | 51(12.2) | 368(87.8) |
| 12. | Set yourself up in a relationship to be rejected? | 52(12.4) | 367(87.6) |
| 13. | Abused prescription drugs/medication? | 46(11.0) | 373(89.0) |
| 14. | Distanced yourself from God as punishment? | 39(9.3) | 380(90.7) |
| 15. | Engaged in emotionally abusive relationships? | 44(10.5) | 375(89.5) |
| 16. | Engaged in sexually abusive relationships, such as being beaten or maltreated by a girlfriend or boyfriend | 45(10.7) | 374(89.3) |
| 17. | Lost a job on purpose? | 45(10.7) | 373(89.0) |
| 18. | Attempted suicide or wanted to kill yourself? | 24(5.7) | 395(94.3) |
| 19. | Did you inflict an injury on yourself on purpose? | 41(9.8) | 378(90.2) |
| 20. | Tortured yourself with self-defeating thoughts? | 46(11.0) | 373(89.0) |
| 21. | Starved yourself to hurt yourself (Refused to eat food to hurt yourself)? | 51(12.2) | 368(87.8) |
| 22. | Abused laxatives (drugs that make someone vomit) to hurt yourself? | 26(6.2) | 393(93.8) |
|  | **Average %** | **11.4** | **88.6** |

**Note: 0-39 % = low proportion; 40-59 % = moderate proportion; 60-79 % = High proportion; ≥ 80 % = very high proportion.**

The most prevalent forms of NSSI reported were overdosing on drugs (19.8%), preventing wound healing (15.5%), intentionally worsening medical conditions by skipping prescribed medications (15.0%), reckless driving of parents’ car or motorcycle (13.8%), and self-hitting (13.8%). Other commonly reported behaviors included cutting oneself (12.2%), starving oneself as a form of self-punishment (12.2%), and entering relationships with the intent of rejection (12.4%).

Less frequent behaviors included burning oneself (7.9%), abuse of alcohol (8.8%), abuse of prescription medication (11.0%), and engagement in emotionally (10.5%) or sexually abusive relationships (10.7%). The least reported behaviours were attempted suicide or suicidal ideation (5.7%) and abuse of laxatives for self-harm (6.2%).

Table 3 presents the mean scores on the PHQ-9 items for in-school adolescents in Gboko LGA, Benue State (n = 419). The overall mean PHQ-9 score was 11.41 (SD = 10.35), indicating a moderate level of depressive symptoms in the study population. Item-level means showed that the most frequently reported symptoms were trouble concentrating 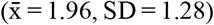, psychomotor changes or restlessness 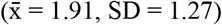, feelings of worthlessness or failure 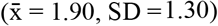, and appetite disturbance 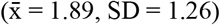. Fatigue or low energy 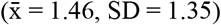 was also moderately reported. By contrast, lower mean scores were observed for anhedonia 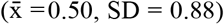, depressed mood 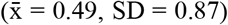, and sleep disturbance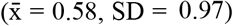. Suicidal ideation 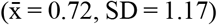 was considered less frequently but remains clinically significant.

**Table 3:** Mean and Standard Deviation Showing Level of Depression among In-school Adolescents in Gboko LGA, Benue State (n = 419)

| S/N | PHQ-9 Items | $\bar{x}$ | SD |
| --- | --- | --- | --- |
|  | Over the last 2 weeks, how often have you been bothered by any of the following: |  |  |
| 1. | little interest or pleasure in doing things | 0.5 | 0.88 |
| 2. | Feeling down, depressed, or hopeless | 0.49 | 0.87 |
| 3. | Trouble falling or staying asleep, or sleeping too much | 0.58 | 0.97 |
| 4. | Feeling tired or having little energy | 1.46 | 1.35 |
| 5. | Poor appetite or overeating | 1.89 | 1.26 |
| 6. | Feeling bad about yourself or that you are a failure or have let yourself or your family down | 1.9 | 1.3 |
| 7. | Trouble concentrating on things, such as reading the newspaper or watching television | 1.96 | 1.28 |
| 8. | Moving or speaking so slowly that other people could have noticed. Or the opposite, being so fidgety or restless that you have been moving around a lot more than usual | 1.91 | 1.27 |
| 9. | Thoughts that you would be better off dead, or of hurting yourself | 0.72 | 1.17 |
|  | **Total PHQ-9 Score** | **11.41** | **10.35** |

**PHQ-9 = Nine-item Patient Health Questionnaire $\bar{x}$ =Mean; SD=Standard Deviation**

**Table 4:** Mean and Standard Deviation Showing Level of Anxiety among In-school Adolescents in Gboko LGA, Benue State (n-419)

| S/N | GAD-7 Items | $\bar{x}$ | SD |
| --- | --- | --- | --- |
|  | Over the last 2 weeks, how often have you been bothered by the following problems? |  |  |
| 1. | Feeling nervous, anxious, or on edge | 0.32 | 0.75 |
| 2. | Not being able to stop or control worrying | 0.46 | 0.84 |
| 3. | Worrying too much about different things | 0.6 | 0.94 |
| 4. | Trouble relaxing | 0.7 | 1.09 |
| 5. | Being so restless that it is hard to sit still | 1.27 | 1.34 |
| 6. | Becoming easily annoyed or irritable | 1.95 | 1.28 |
| 7. | Feeling afraid, as if something awful might happen | 2.02 | 1.24 |
|  | **Total GAD - 7 Score** | **7.32** | **7.48** |

Note. GAD – 7 = 7-item Generalised Anxiety Disorder, Scoring Protocol for interpreting Anxiety level/severity, 0-4 = Minimal Anxiety, 5-9= Mild Anxiety, 10-14 = Moderate Anxiety, 15-21 = Severe Anxiety

Therefore, the result showed that the in-school adolescents in Gboko LGA had a moderate level of depression 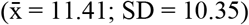. The results suggest that in-school adolescents experienced depression symptoms considerably in Gboko LGA, Benue State.

Results in Table 3 show that in-school adolescents in Gboko LGA had a mild level of anxiety 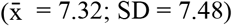. This result suggests that in-school adolescents were mildly anxious in Gboko LGA, Benue State.

## Discussion of Findings

### Proportion of in-school adolescents who engage in NSSI behaviours

Findings in Table 1 show that in-school adolescents had a low proportion of engagement in NSSI behaviour. The finding was somewhat expected, considering the strong familial and community ties often observed in Nigerian cultural settings, which may provide protective support against such behaviours. This finding aligns with studies conducted in other African contexts, such as one by Naidoo (2019), which reported low NSSI prevalence in school-aged adolescents due to robust family bonds and communal values that discourage self-injury. However, it contrasts with findings from Western countries, such as those by Klonsky, Victor, and Saffer (2014), where NSSI prevalence is higher, often attributed to increased individualism, mental health stigma, and less emphasis on collective family support. The agreement with African-based studies could stem from similar cultural and societal influences, while the contrast with Western studies likely reflects differences in cultural norms, mental health awareness, and access to mental health resources. The relatively low prevalence in this study may also be due to underreporting, influenced by stigma surrounding self-injury and mental health issues in the Nigerian context. This highlights the need for culturally tailored interventions and improved mental health awareness to ensure accurate identification and support for at-risk adolescents.

### Level of depression among in-school adolescents

Findings in Table 2 revealed that in-school adolescents had a moderate level of depression in Gboko LGA, Benue State. The finding was somewhat expected, given the numerous stressors adolescents face during their developmental years, including academic pressure, social challenges, and family expectations. This result aligns with studies conducted in other parts of Nigeria, such as the work by Adewuya et al. (2007), which also found moderate levels of depression among Nigerian adolescents, likely due to social and environmental stressors. The findings also agree with a more recent study by Nkporbu and Alex-Hart (2022), which highlighted a similar prevalence of depressive symptoms among Nigerian adolescents, further pointing to the growing mental health challenges within this demographic. However, the study contrasts with findings of Thapar, Collishaw, Pine, and Thapar (2012) who reported higher levels of depression, possibly due to greater individualism, less robust family structures, and a higher prevalence of mental health issues in high-income settings. The contrast may reflect the more communal nature of Nigerian society, where familial support systems and cultural attitudes toward mental health might buffer adolescents against more severe depressive symptoms. The moderate levels of depression observed in this study could also result from the stigmatization of mental health issues in Nigeria, leading adolescents to underreport more severe symptoms.

### Level of anxiety among in-school adolescents

Furthermore, findings in Table 3 show that in-school adolescents had a mild level of anxiety in Gboko LGA, Benue State. The finding was expected given the developmental stage of adolescence, which is often characterized by moderate emotional and psychological stressors, such as academic pressure, peer relationships, and family expectations. This result aligns with studies conducted in similar settings, such as Adewuya, Ola, and Adewumi (2007), which reported mild-to-moderate anxiety levels among Nigerian adolescents, attributing it to protective factors like family support and cultural norms that promote communal coping mechanisms. However, it contrasts with findings from studies in high-income countries, such as Costello, Egger, and Angold (2003), which observed higher levels of anxiety among adolescents, likely due to different sociocultural dynamics, greater individualism, and the absence of communal support systems. The agreement with Nigerian-based studies reflects the role of family and cultural factors in mitigating anxiety, while the contrast with Western studies may highlight differences in mental health awareness, stigma, and the availability of mental health services. The mild level of anxiety observed in this study might also be influenced by underreporting, as mental health issues are often stigmatised in Nigerian society, leading adolescents to downplay their symptoms.

## Conclusion

This study investigated the prevalence of non-suicidal self-injury (NSSI), depression, and anxiety among in-school adolescents in Gboko Local Government Area of Benue State, Nigeria. The findings revealed a relatively low engagement in NSSI behaviours, a moderate level of depression, and a mild level of anxiety among the participants. These outcomes reflect the influence of strong familial and communal ties prevalent in Nigerian society, which appear to serve as protective factors against severe mental health challenges. The low prevalence of NSSI aligns with similar findings in other African contexts, suggesting that cultural norms and collective support systems may discourage self-injurious behaviours. However, the contrast with higher NSSI rates reported in Western settings underscores the impact of sociocultural differences, particularly regarding individualism and mental health stigma. Moderate levels of depression among the adolescents were consistent with prior Nigerian studies, indicating that while protective factors exist, adolescents still face considerable psychosocial stressors. Similarly, the mild levels of anxiety observed suggest that while emotional challenges are present, they may be buffered by communal coping mechanisms and familial support. Nonetheless, the possibility of underreporting due to stigma surrounding mental health issues in Nigeria cannot be overlooked. This highlights the urgent need for culturally sensitive mental health interventions, increased awareness, and destigmatization efforts to ensure accurate assessment and effective support for adolescents experiencing psychological distress. Conclusively, the study underscores the complex interplay between cultural context and adolescent mental health, and calls for targeted strategies that respect and leverage local values while addressing emerging mental health needs.

### Recommendations

Based on the findings and conclusion of the study, the following recommendations are made:

1. Schools should include mental health education as part of their regular curriculum to promote awareness of depression, anxiety, and NSSI behaviours. This should include teaching adolescents coping strategies, stress management techniques, and how to seek help when experiencing emotional challenges.
2. Public health interventions should focus on enhancing parental attachment and involvement in adolescents’ lives. Workshops and community programs can be organized to teach parents effective communication, emotional support strategies, and how to recognize signs of distress in their children.
3. Schools should provide access to trained counsellors who can offer support and interventions for students struggling with mental health issues. These services should include confidential counselling sessions, peer-support programs, and regular mental health assessments to identify at-risk students early.

## Data Availability

All data produced in the present study are available upon reasonable request to the authors

